# Slow Motion Analysis of Repetitive Tapping (SMART) test: measuring bradykinesia in recently diagnosed Parkinson’s disease and idiopathic anosmia

**DOI:** 10.1101/2021.03.24.21254234

**Authors:** C. Simonet, MA. Galmes, C. Lambert, RN. Rees, T. Haque, JP. Bestwick, AJ. Lees, A. Schrag, AJ. Noyce

**Author notes:** **Corresponding author:** Dr Alastair Noyce., Preventive Neurology Unit, Wolfson Institute of Preventive Medicine, Barts and the London School of Medicine and Dentistry, Queen Mary University of London, London, United Kingdom.

## Abstract

**Background:** Bradykinesia is the defining motor feature of Parkinson’s disease (PD). There are limitations to its assessment using standard clinical rating scales, especially in the early stages of PD when a floor effect may be observed.

**Objectives:** To develop a quantitative method to track repetitive finger tapping movements and to compare people in the early stages of PD, healthy controls, and individuals with idiopathic anosmia.

**Methods:** This was a cross-sectional study of 99 participants (early-stage PD=26, controls=64, idiopathic anosmia=9). For each participant, repetitive finger tapping was recorded over 20 seconds using a smartphone at 240 frames per second. Three parameters were extracted from videos: amplitude between fingers, frequency (number of taps per second), and velocity (distance travelled per second). Clinical assessment was based on the motor section of MDS-UPDRS.

**Results:** People in the early stage of PD performed the task with slower velocity (*p*<0.001) and with greater decrement in frequency than controls (*p=*0.003). The combination of slower velocity and greater decrement in frequency obtained the best accuracy to separate early-stage PD from controls based on metric thresholds alone (AUC = 0.88). Individuals with anosmia exhibited slower velocity (*p=*0.001) and smaller amplitude (*p<*0.001) compared with controls.

**Conclusions:** We present a new simple method to detect early motor dysfunction in PD. Mean tap velocity appeared to be the best parameter to differentiate patients with PD from controls. Patients with anosmia also showed detectable differences in motor performance compared with controls which may be important indication of the prodromal phase of PD.

## 1. INTRODUCTION

The diagnosis of Parkinson’s disease (PD) depends on the detection of bradykinesia *[1–4]* but in the early stages of disease this may not be easy to see. Bradykinesia is defined as slow velocity of movement but is often seen in combination with other abnormalities of movement. These include hypokinesia (reduced amplitude), akinesia (slow initiation contributing to changes in sequence rhythm) and decrement, otherwise known as “sequence effect”, where there is progressive reduction in the velocity or amplitude with repeated movements. These abnormalities of movement can be detected in gait, arm swing, facial expression and handwriting. Many of the common rating scales for PD assess these features, and others, in combination *[5,6]*.

Bradykinesia is elicited clinically by sequential finger or foot tapping and can be scored using the motor section of the Movement Disorders Society-Unified Parkinson’s Disease Rating Scale (MDS-UPDRS-III) *[7]*. For diagnosis, assessment of the whole clinical picture is necessary and reliance should not be placed exclusively on finger tapping *[8]*. While the MDS-UPDRS-III is a useful research scale, the integers prevent adequate detection of subtle motor changes. In particular, bradykinesia-related sub-scores have imperfect interrater reliability *[9]*. Part of this variability may be due to the mixed definition of bradykinesia used by the MDS-UPDRS-III, assigning equal weighting to speed, amplitude, and rhythm with no provision to sub-classify them further. This is of particular relevance to the stage of PD close to diagnosis (based on motor criteria), where current questionnaires and scales may be insufficiently sensitive to detect change, reflecting the need for more accurate and specific measures to detect subtle motor dysfunction *[10]*.

Attempts to develop quantitative measurements of bradykinesia that would be useful in clinical practice began fifty years ago, but many devices are too insensitive or cumbersome for routine clinical use *[11]*. Wearable sensors have shown promise *[12]* but although these offer the potential of 24-hour monitoring, there are limitations such as lack of context to movement, interference with the natural range of movement and cost. There is also a lack of consensus about which derived metrics are best to assess the subtle motor changes in early stage disease *[13]*. This study aims to provide proof of concept that motion capture using a smart phone could assess different elements of bradykinesia which may be sensitive to change in early PD.

## 2. MATERIALS AND METHODS

This was a cross-sectional, case-control study in which the main aim was to design a test to objectively quantify early patterns of motor dysfunction in PD. Repetitive finger tapping movements were recorded using an ordinary smartphone (iPhone X®) with slow motion video capture. Slow Motion Analysis of Repetitive Tapping (SMART) test results were compared in patients with early PD (less than two years since diagnosis), healthy controls and patients with idiopathic anosmia. Parameters derived from the SMART test were correlated with clinical ratings scored from the gold standard of assessment for PD, the MDS-UPDRS-III *[14]*.

### Participants

All the patients with PD fulfilled the UK Queen Square Brain Bank criteria *[1]*. Exclusion criteria included disease duration (defined as time from diagnosis on motor criteria) of more than two years, and any comorbidities that could interfere with performance of the task, such as arthritis, previous stroke, and dementia. Healthy controls were also excluded if they scored more than 6 on the MDS-UPDRS-III (a cut off for subthreshold parkinsonism *[7]*). Cases with PD were recruited from two studies; the East London Parkinson’s disease (ELPD) project based at Barts Health NHS Trust and Quantitative MRI for Anatomical Phenotyping in Parkinson’s disease (qMAP-PD) study based at the Institute of Neurology, University College London. Controls were recruited from the PREDICT-PD study (www.predictpd.com) *[15]* and qMAP-PD study (https://gtr.ukri.org/projects?ref=MR%2FR006504%2F1). Patients with anosmia were recruited from the PREDICT-PD study, after referral from specialist ENT clinics, where nasal endoscopy and imaging, had revealed no identifiable cause of smell loss. Assessments were carried out between October 2018 and December 2019 and all patients gave informed written consent to the study.

### Assessment

Finger tapping was performed following the same standardised instructions that are used when administering the MDS-UPDRS-III (see supplementary material). Movements were recorded over 20 seconds using a smartphone at 240 frames per second (slow motion capture). In order to facilitate finger recognition by the software, we asked participants to tap their index finger on the thumb ‘as fast and as wide’ as they could while making a fist with the remaining three fingers (**Figure 1**). Participants were instructed to not rotate and move the arm during the task with the purpose of capturing the angle at the metacarpal-phalangeal joints between index finger and thumb. Patients were asked to stop taking any dopaminergic medication at least 12 hours before the assessment. In order to compare their performance under ‘on’ and ‘off’ medication they were tested again having been taken their regular dopaminergic medication.

**Figure 1.**
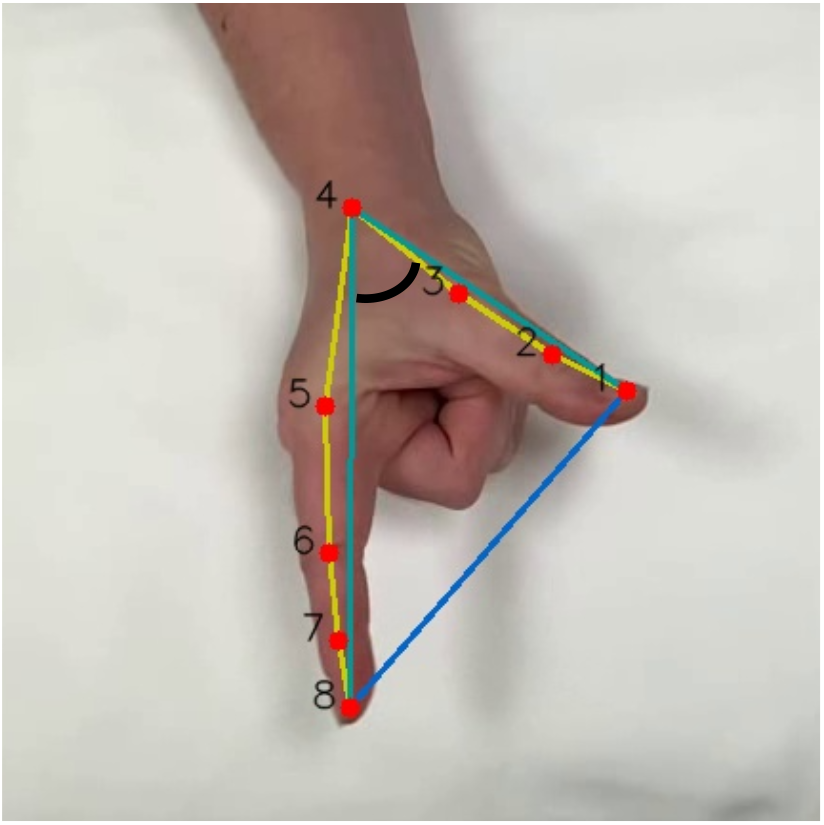
Hand detection: 8 key landmarks across the first and the second finger (red). Angle between 1,4,8 key landmarks (black). Extrapolated amplitude between point 1 and 8 (blue).

### Video analysis

We created a convolutional neural network (CNN), which was built using PyTorch 1.6.0 *[16]*, to detect the shape of the hand in the video. This enabled the tracking of movement of the hand during the tapping task. We also built a 2D CNN which was trained to detect 8 key landmarks of the index finger and the thumb which were then tracked over time (**Figure 1**). Videos were resized and rotated for standardisation. The ‘pre-processing’ stage was carried out using OpenCV library *[17]*. Twenty frames were randomly extracted from each video and used as a dataset to train the CNN, making a total of 3934 frames in the initial dataset. The architecture of the CNN was divided into 8 blocks of 2D convolutional layers followed by a batch normalisation, 4 pooling layers and a final 3 fully connected layers, using the ReLU activation function. To measure the accuracy, we computed the deviation as the Euclidean distance between manual and predicted landmarks on the test dataset. We achieved an average deviation of 11.3 ± 8.6 pixels on the final 606 x 1080 images (i.e. an average error of 0.9%).

Once the training was completed, videos were processed using the CNN frame by frame to extract the predicted anatomical landmarks. After the position of the key landmarks had been predicted, the distances between the distal portion of the index finger and the thumb were calculated (**Figure 1**). Although normalising the amplitude allowed comparison between samples, the absolute amplitude was needed to calculate the initial and mean amplitude (fully separating the finger from the thumb for one individual is not the same as for another individual), as well as the change in amplitude over time. To overcome this limitation, the angle formed between the distal part of the index finger and the thumb and the key landmark corresponding to the metacarpal joint was computed (i.e. the angle formed between landmarks 1-4-8 in **Figure 1**) to mitigate the need for an external reference to normalise amplitude.

Maximum amplitude peaks were detected for each tap and linear regression models were fitted to those signal peaks. Frequency was measured as the number of taps per second. Velocities were calculated as the change rate of the normalised signal, and a similar process was applied to obtain the peaks of maximum velocities along time. All the signal processing was done using SciPy *[18]* and NumPy libraries *[19]*.

### Statistical analysis

Three kinetic parameters were extracted to be used in the statistical analysis: amplitude (angle formed between index finger and thumb), frequency (number of taps per second) and velocity (distance travelled per second extracted from the derivative of the amplitude). For each parameter, the mean, coefficient variation (CV) (standard deviation divided by the mean), and slope (from regression of time against each parameter) was calculated.

Normality of the data was assessed using the D’Agostino test. Quantitative data was presented as the median and interquartile range (IQR) for non-parametric data and the mean and standard deviation (SD) for parametric data. Mann Whitney U tests, t-tests, and Welch’s t-tests (two-tailed) were used to compare test parameters between patients and controls, as appropriate. Linear regression was used to determine whether movement parameters derived from finger tapping (dependent variables) were influenced by age and time since the last dose of levodopa. Logistic regression was performed to examine whether test parameters were associated with binomial factors such as gender and handedness. Receiver operator characteristic (ROC) curves were drawn to find the optimal cut off value with the best combination of sensitivity and specificity for SMART test parameters separately and in combination. Spearman’s correlation coefficient was used to correlate SMART test variables with finger-tapping sub-scores from the MDS-UPDRS-III. Since multiple hypothesis tests were run, one for each component of the test parameters (mean, CV, and slope), a more stringent cut-off for the level of significance (*p<*0.005, Bonferroni corrected for nine hypothesis tests) was selected to ensure robustness of results and avoid false positives (i.e. type I error). Data analysis was carried out using STATA V.13 (StataCorp, College Station, TX).

## 3. RESULTS

Two hundred and ninety-four videos were analysed (99 recordings for the right and left hands for all participants, and recordings for the right and left hands of 24 patients during ‘on’ and ‘off’ medication recordings). There was no significant difference in the derived motor metrics between the dominant and non-dominant hands in the control group, therefore we focused further analysis on results derived from the dominant hand in the controls and anosmic cohorts. Since PD is, by definition, associated with asymmetric onset of motor signs and the patients were all in early disease stage, the most severely affected side was used for the analysis. The identification of the most affected side was based on the side with the worst finger-tapping sub-scores in the MDS-UPDRS-III.

### Early PD

#### Clinical and demographic information

Twenty-six patients with early PD and 30 controls were included in the first analysis. The other 34 controls were on average much older than the PD patients and were excluded to make both groups more comparable (PD: 59.60 years, SD 10.88 vs Control: 63.81 years, SD 7.21, p-value=0.060). Compared with controls, PD cases were more likely to be male (65.38% vs 36.67%, *p*=0.030). All patients had a disease duration of less than two years (median 0.75 years, IQR 0.5-1.2) and were taking levodopa. The mean MDS-UPDRS-III score was 21.2 ± 8.3 points (range 11–47). Most of the patients exhibited abnormal finger-tapping to a slight-mild degree (12 patients scored 1 and 12 patients scored 2 in the MDS-UPDRS-III sub-score). One patient was found to had normal finger-tapping and another one showed a moderately (score 3) abnormal finger-tapping performance. **Table 1** summarises the clinical and demographic information of both groups. SMART test parameters were not associated with age, gender or handedness (*p*>0.05 for all parameters).

**Table 1.** Demographic and clinical data.

|  | Control<sup>1</sup><br>(n=30)<br>-from control<sup>2</sup>- | PD<br>(n=26) | Control<sup>2</sup><br>(n=64) | Anosmia<br>(n=9) |
| --- | --- | --- | --- | --- |
| Age, years (SD) | 63.81 (7.21) | 59.60 (10.88) | 69.19 (7.68) | 70.94 (8.17) |
| Gender, male: female | 11:19 | 17:9 | 26:38 | 7:2 |
| Median years since PD diagnosis (IQR) | NA | 0.75 (0.5-1.2) | NA | NA |
| Last dose of LD, median hours (IQR) | NA | 16.6 (15-21) | NA | NA |
| Median MDS-UPDRS-III score (IQR) | 1 (0-2) | 20 (15-26) | 1.5 (0-3) | 1 (1-3) |
| Median MDS-UPDRS-III score worsening<br>(on-off medication) | NA | 25% (13%-<br>61%) | NA | NA |
| Visible tremor during task | 0 | 11 | 0 | 0 |
| FT sub-score (MDS-UPDRS-III) |  |  |  |  |
| 0 | 30 | 1 | 63 | 6 |
| 1 | 0 | 12 | 1 | 2 |
| 2 | 0 | 12 | 0 | 1 |
| 3 | 0 | 1 | 0 | 0 |
| 4 | 0 | 0 | 0 | 0 |

\*Finger tapping (FT) sub-score in the MDS-UPDRS-III: 0-normal, 1- slight, 2-mild, 3-moderate, 4-severe. IQR: interquartile range, SD: standard deviation, NA: not applicable. Overall, 64 controls were included. Group (1): 30 out of 64 were extracted to compare with PD. Group (2): overall control group used for comparison with anosmia.

#### SMART scores

Patients with PD performed repetitive finger tapping with slower velocity (PD: 1.20 degrees/s, 95% CI 1.02 to 1.38 vs Control: 1.63 degrees/s, 95% CI 1.44 to 1.81 *p<*0.001) but similar amplitude to controls with wider confidence interval (CI) and overlap between both groups (PD: 27.08, 95% CI 22.49 to 31.67 vs Control: 31.10 degrees, 95% CI 26.91 to 35.28, *p*=0.189). There was some evidence that patients with PD displayed greater variability (higher CV) in frequency (PD: 0.18, 95% CI 0.13 to 0.22 vs Control: 0.11, 95% CI 0.08 to 0.14, *p*=0.007) and more so in velocity compared with controls (PD: 0.31, 95% CI 0.27 to 0.34 vs Control: 0.20, 95% CI 0.15 to 0.25 *p<*0.001). There was also more evident decrement of frequency in patients than controls (PD: −0.02, 95% CI −0.03 to 0.01 vs Control: −0.002, 95% CI −0.01 to 0.007, *p*=0.003) (**Table 2**).

**Table 2.** Test parameter comparison and ROC analysis between PD and controls.

|  | Controls | PD | *p* value |
| --- | --- | --- | --- |
| **Amplitude** Mean | 31.10 (26.91 to 35.28) | 27.08 (22.49 to 31.67) | 0.189 |
| CV | 0.18 (0.14 to 0.23) | 0.21 (0.17 to 0.25) | 0.447 |
| Slope | -0.42 (-0.58 to 0.27) | -0.39 (-0.62 to -0.17) | 0.817 |
| **Frequency** Mean | 3.18 (2.84 to 3.53) | 2.63 (2.29 to 2.98) | 0.017 |
| CV | 0.11 (0.08 to 0.14) | 0.18 (0.13 to 0.22) | 0.007 |
| Slope | -.002 (-0.01 to 0.007) | -0.021 (-0.03 to 0.01) | 0.003 |
| **Velocity** Mean | 1.63 (1.44 to 1.81) | 1.20 (1.02 to 1.38) | <0.001 |
| CV | 0.20 (0.15 to 0.25) | 0.31 (0.27 to 0.34) | <0.001 |
| Slope | -0.06 (-0.08 to -0.04) | -0.07 (-0.08 to -0.05) | 0.662 |

All parameters presented with 95% coefficient interval (CI). CV: coefficient variation. Amplitude: degrees. Frequency: taps/sec. Velocity: degrees/sec. P-value: Welch's t-tests (two-tailed) except for frequency were Two-sample Wilcoxon rank-sum (Mann-Whitney) test was used

Action tremor was visible in eleven patients. To prevent over estimation of a high frequency caused by tremor, when two consecutive peaks of amplitude appeared without reaching the baseline amplitude 0, it was interpreted as a tremor. The highest peak was selected to avoid under estimation of the amplitude (**Figure 4**). In some patients a re-emergent action tremor was seen with the tremor occurring after a finite period (latency) from the time the patient started the finger tapping task (illustrated in **Figure 4**).

#### Diagnostic accuracy

Velocity offered the best discriminatory power with 84.62% sensitivity for 73.33% specificity and an AUC of 0.81 (95% CI 0.69 to 0.93). The CV of frequency also showed good discrimination with 80.77% sensitivity for 70% specificity and an AUC of 0.75 (95% CI 0.62 to 0.88). Combining both parameters (velocity and the CV of frequency) meant that the specificity improved to 86.67% with sensitivity remaining the same (AUC 0.83; 95% CI 0.72 to 0.95). The slope of frequency was able to distinguish between groups with a moderate accuracy (AUC 0.72; 95% CI 0.59 to 0.86), but when it was combined with velocity the discriminatory power improved, yielding a sensitivity of 80.77% for 83.33% specificity (AUC 0.88, 95% CI 0.78 to 0 0.97). In the same way, when decrement of frequency was combined with CV velocity, both parameters also reached a high accuracy (AUC 0.85; 95% CI 0.74 to 0.95) with 80.77% sensitivity for 85% specificity (**Table 3 and Figure 2**).

**Table 3.**
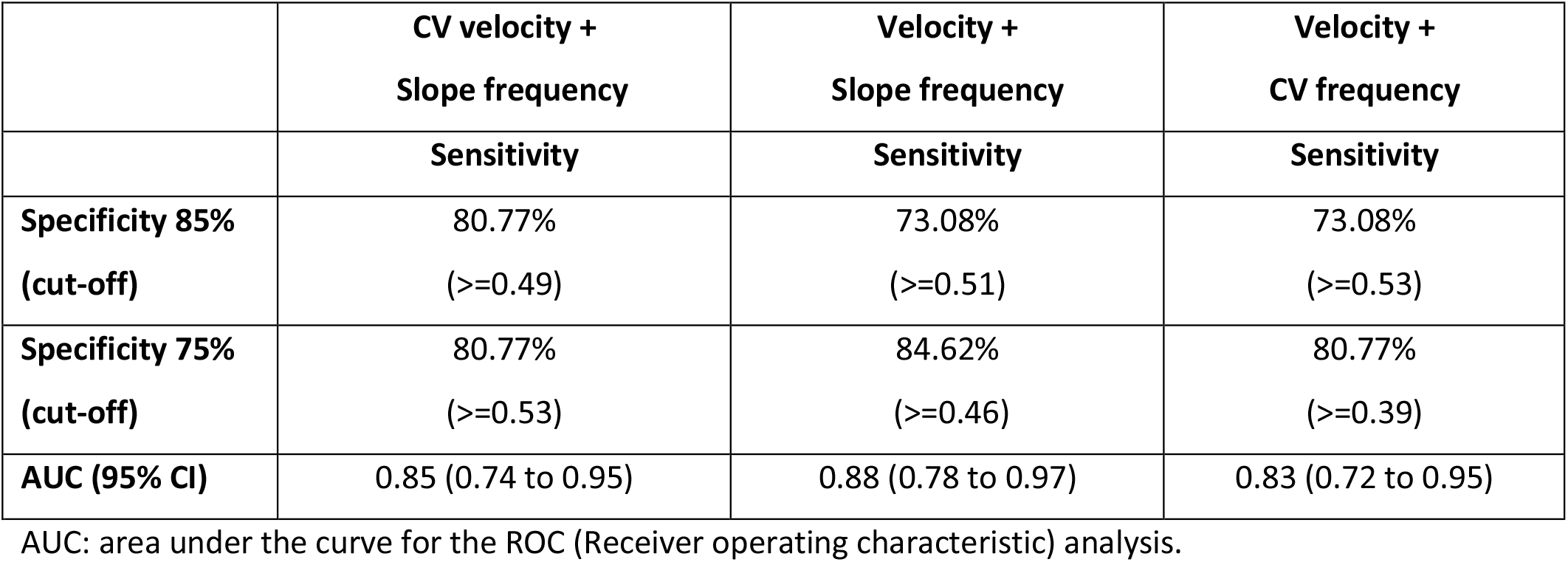
ROC analysis between PD and control group.

|  | **CV velocity +**<br>**Slope frequency** | **Velocity +**<br>**Slope frequency** | **Velocity +**<br>**CV frequency** |
| --- | --- | --- | --- |
|  | **Sensitivity** | **Sensitivity** | **Sensitivity** |
| **Specificity 85%**<br>**(cut-off)** | 80.77%<br>($\geq 0.49$) | 73.08%<br>($\geq 0.51$) | 73.08%<br>($\geq 0.53$) |
| **Specificity 75%**<br>**(cut-off)** | 80.77%<br>($\geq 0.53$) | 84.62%<br>($\geq 0.46$) | 80.77%<br>($\geq 0.39$) |
| **AUC (95% CI)** | 0.85 (0.74 to 0.95) | 0.88 (0.78 to 0.97) | 0.83 (0.72 to 0.95) |

AUC: area under the curve for the ROC (Receiver operating characteristic) analysis.

**Figure 2.**
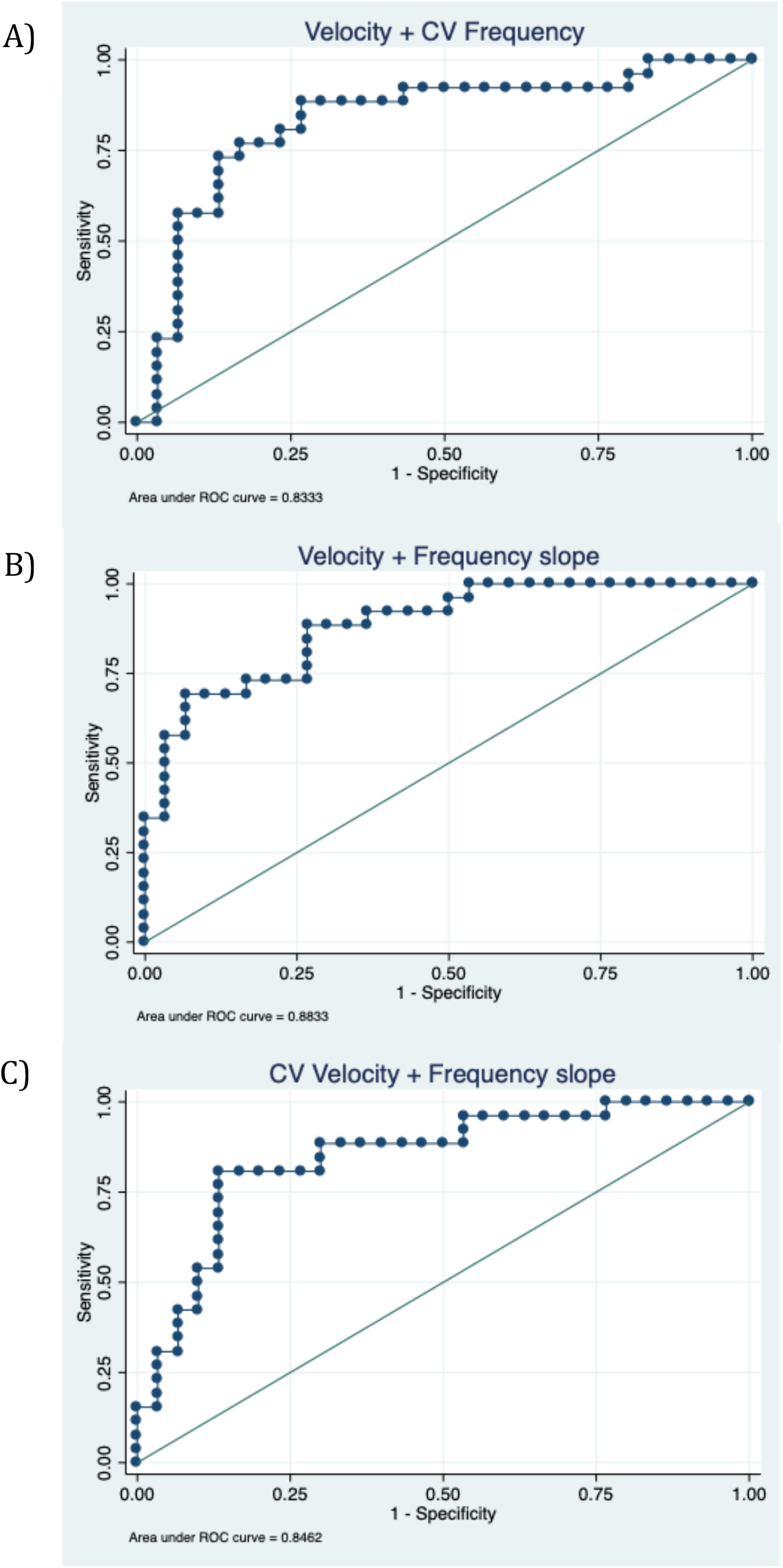
Receiver operator characteristic (ROC) curves for the best parameter combination to distinguish patients with PD and controls. A) Velocity and CV frequency (AUC 0.83; 95% CI 0.72 to 0.95), B) Velocity and frequency slope (AUC 0.88, 95% CI 0.78 to 0 0.97), C) CV velocity and frequency slope (AUC 0.85; 95% CI 0.74 to 0.95).

#### Clinical correlation

Correlations between the three SMART test parameters and finger tapping sub-scores of the MDS-UPDRS-III were examined in patients with PD (for sub-scores definition see **Table 1** in the supplementary material). All PD patients except two scored between 1 (slight degree) and 2 (mild degree) in the MDS-UPDRS-III sub-score. In order to avoid the two patients scoring 0 (normal degree) and 3 (moderate degree) influencing the correlation curves (Figure 1 in supplementary material), they were excluded from the main correlation analysis. Thus, the mean amplitude was found to have the highest correlation with finger tapping score (r= −0.49, *p=*0.003) followed by velocity (r= −0.43, *p=*0.016), whereas there was no correlation with frequency. For more detailed information about the correlations explored see **Table 2** and **Figure 1** in the supplementary material.

#### ‘On’ and ‘off’ medication

For 24 of the patients with PD, it was possible to assess them both ‘on’ and ‘off’ dopaminergic medication. All participants except one experienced a worsening in their MDS-UPDRS-III total score with a median of 25% increase in scores from ‘on’ to ‘off’ medication. In contrast, medication did not change MDS-UPDRS finger tapping sub-score in more than a half of patients (62.50%). Approximately seven patients with PD experienced a worsening in their FT score (from 0 -normal-to 1 -slight-) and in 2 patients their score improved by 1 point. From SMART recordings, no significant differences were found in any of the parameters (amplitude, frequency, and velocity) between ‘on’ and ‘off’ medication.

### Idiopathic anosmia group

#### Clinical and demographic information

Patients with idiopathic anosmia were older on average than patients with PD, with similar age to the control group (Anosmia: 70.94 years SD 8.17 vs Control: 69.19 years SD 7.68, *p=* 0.581) and were therefore compared with the full number of controls. Mean duration since diagnosis of anosmia was 5.25 years (SD 4.65 years). In total, nine patients with idiopathic anosmia and 64 controls were included in a secondary analysis. There was a higher proportion of males in the anosmia group compared to controls (Anosmia: 77.78% males vs Control: 40.62% male, *p*=0.069). The median motor score on the MDS-UPDRS-III was 1 (IQR= 0-3) and no patients met the diagnostic criteria for PD. However, one individual, who scored 10 on the MDS-UPDRS-III, was classified as having sub-threshold parkinsonism based on MDS Task Force criteria (cut off >6 excluding action tremor) *[7]*. The remaining patients with anosmia scored between 0 and 4 in the MDS-UPDRS-III. Finger-tapping sub-scores in the MDS-UPDRS-III were normal (score = 0) except for two individuals who exhibited slight bradykinesia (score = 1) and one who was scored as having mild bradykinesia (score = 2). **Table 1** summarises the clinical and demographic information of both groups.

#### SMART scores

Individuals with anosmia appeared to have impairment in finger-tapping performance compared with the control group and similar patterns of movement to patients with PD. Individuals with anosmia performed the task with a reduced average amplitude; despite broad ranges there was no overlap between groups (Anosmia: 13.94 degrees, 95% CI 9.19 to 18.69 vs Control: 29.38 degrees, 95% CI 26.87 to 31.89 *p<*0.001) (**Table 4**). Compared with controls, the anosmia group showed a slower velocity over finger tapping task (Anosmia: 0.96 degrees/s, 95% CI 0.64 to 1.27 vs Control: 1.48 degrees/s, 95% CI 1.37 to 1.60 *p<*0.001). Although average frequency was similar between anosmia and controls, there was weak evidence that individuals with anosmia exhibited slightly greater decrement over time compared with controls (*p*=0.059). In contrast to PD, CV of velocity was similar between groups (*p*=0.054).

**Table 4.** Test parameter comparison between Anosmia and controls.

|  | **Controls** | **Anosmia** | **p value** |
| --- | --- | --- | --- |
| **Amplitude** Mean | 29.38 (26.87 to 31.89) | 13.94 (9.19 to 18.69) | <0.001 |
| CV | 0.19 (0.16 to 0.22) | 0.30 (0.20 to 0.40) | 0.009 |
| Slope | -0.39 (-0.49 to -0.29) | -0.23 (-0.49 to -0.03) | 0.243 |
| **Frequency** Mean | 3.05 (2.82 to 3.28) | 3.26 (2.62 to 3.90) | 0.515 |
| CV | 0.13 (0.10 to 0.16) | 0.15 (0.05 to 0.26) | 0.560 |
| Slope | -.002 (-0.01 to 0.005) | -0.020 (-0.04 to -0.003) | 0.059 |
| **Velocity** Mean | 1.48 (1.37 to 1.60) | 0.96 (0.64 to 1.27) | 0.001 |
| CV | 0.21 (0.18 to 0.23) | 0.28 (0.16 to 0.40) | 0.054 |
| Slope | 0.02 (0.02 to 0.03) | 0.01 (-0.004 to 0.03) | 0.369 |

All parameters presented with 95% coefficient interval (CI). CV: coefficient variation, AUC: area under the curve, ROC: Receiver operating characteristic. P-value: Welch's t-tests (two-tailed)

## 4. DISCUSSION

The main aim of the study was to identify subtle but pathological abnormalities in finger tapping in PD which might be difficult to pick up with the ‘naked eye’ but are detectable through slow-motion video capture. Here we present a quantitative metric to create parameters of a repetitive movement which is often abnormal in established PD.

We found that patients with PD had slower finger tapping in line with the etymological definition of bradykinesia (‘*slowness of movement’*). In addition, we found there was significantly greater decrement in frequency of finger tapping. However, we did not find any difference in either mean amplitude or decrement in amplitude using the SMART test. Slowing, interruptions and reduced amplitude of finger tapping are all aspects typically seen in PD and evaluated in the finger tapping component of the MDS-UPDRS-III. Other studies using electronic measures have yielded similar results *[20]*. One explanation for the failure of these measurements to capture reduction in amplitude might be that change in amplitude in PD cases does not follow a linear trend over time. This was seen in many of the plots extracted from time series of PD cases showing a non-linear trend with a ‘*burst’* phenomenon: repetitive cycles of slowing down and becoming smaller followed by a late amplitude increase. In fact, this last augmentation could compensate for the decrement and the average of amplitude over the 20-second task (see PD case example B in **Figure 4**). This rebound pattern could have a proprioceptive origin, suggesting that it might be an early feature before grinding down to a complete halt in more established PD.

In contrast, kinetic parameters (velocity and frequency) were able to distinguish patients from controls with a good accuracy particularly using a combination of both (AUC 0.88). Our findings agreed with some other studies, with velocity and parameter variation found to have a high accuracy (see **Table 5**). In contrast, in a study by Růžička and colleagues, who used a contactless 3D motion capture system to compare 22 patients with 20 controls, amplitude was the best marker *[21]*. Amplitude decrement alone provided an accuracy of 0.87. Since their cases had a longer disease duration (9.3 years) than ours, this might suggest that ‘*sequence effects’* are more apparent later in the disease course.

**Figure 3.**
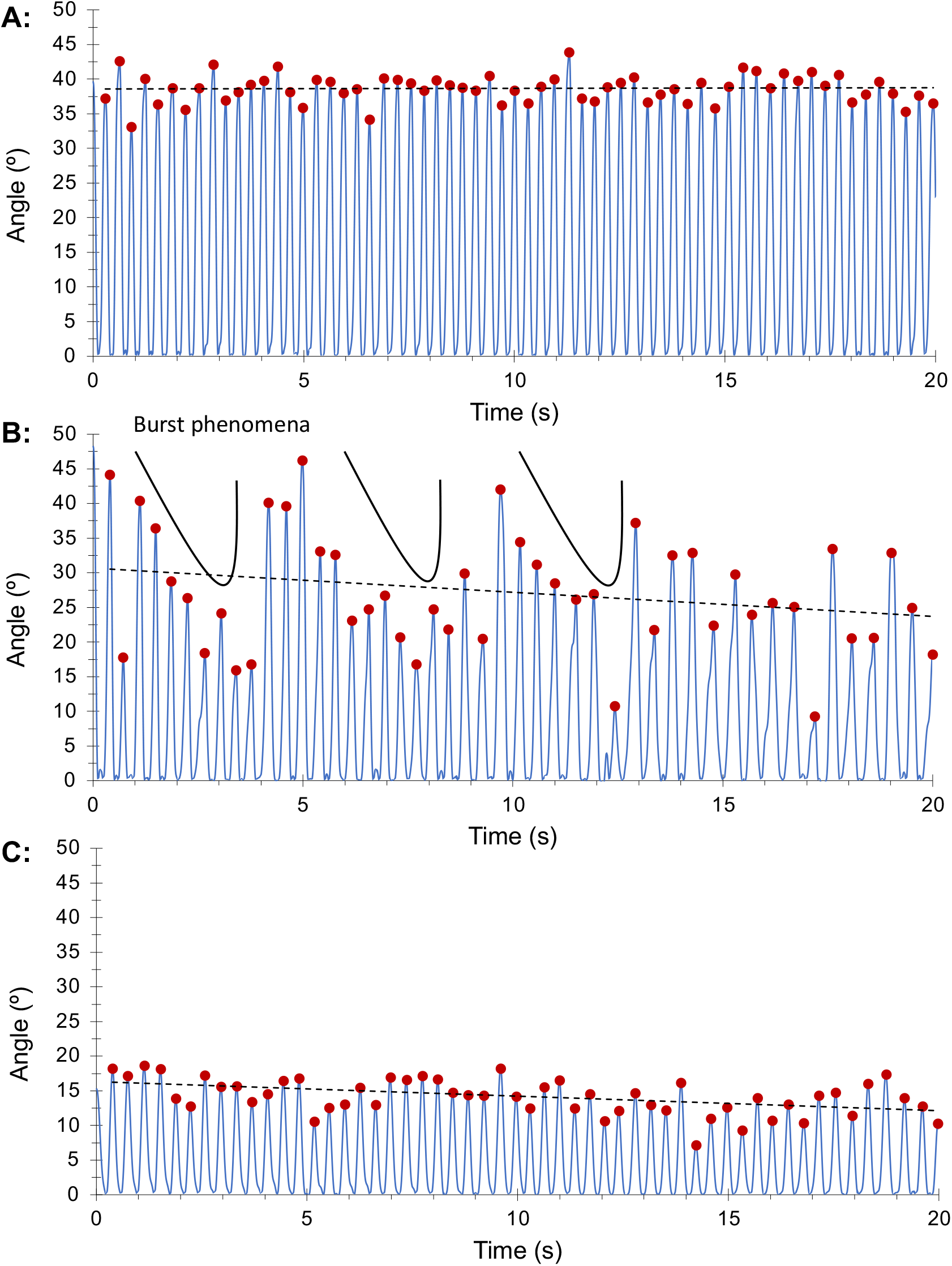
Representative examples of one control (A), PD (B), and anosmia (C). Finger tapping sub-scores (MDS-UPDRS-III): 0 (control and anosmia), 1 (PD). Control: constant amplitude and frequency. PD amplitude with ‘*burst phenomena*’ and frequency decrease. Anosmia: smaller amplitude and frequency. compared with control.

**Figure 4.**
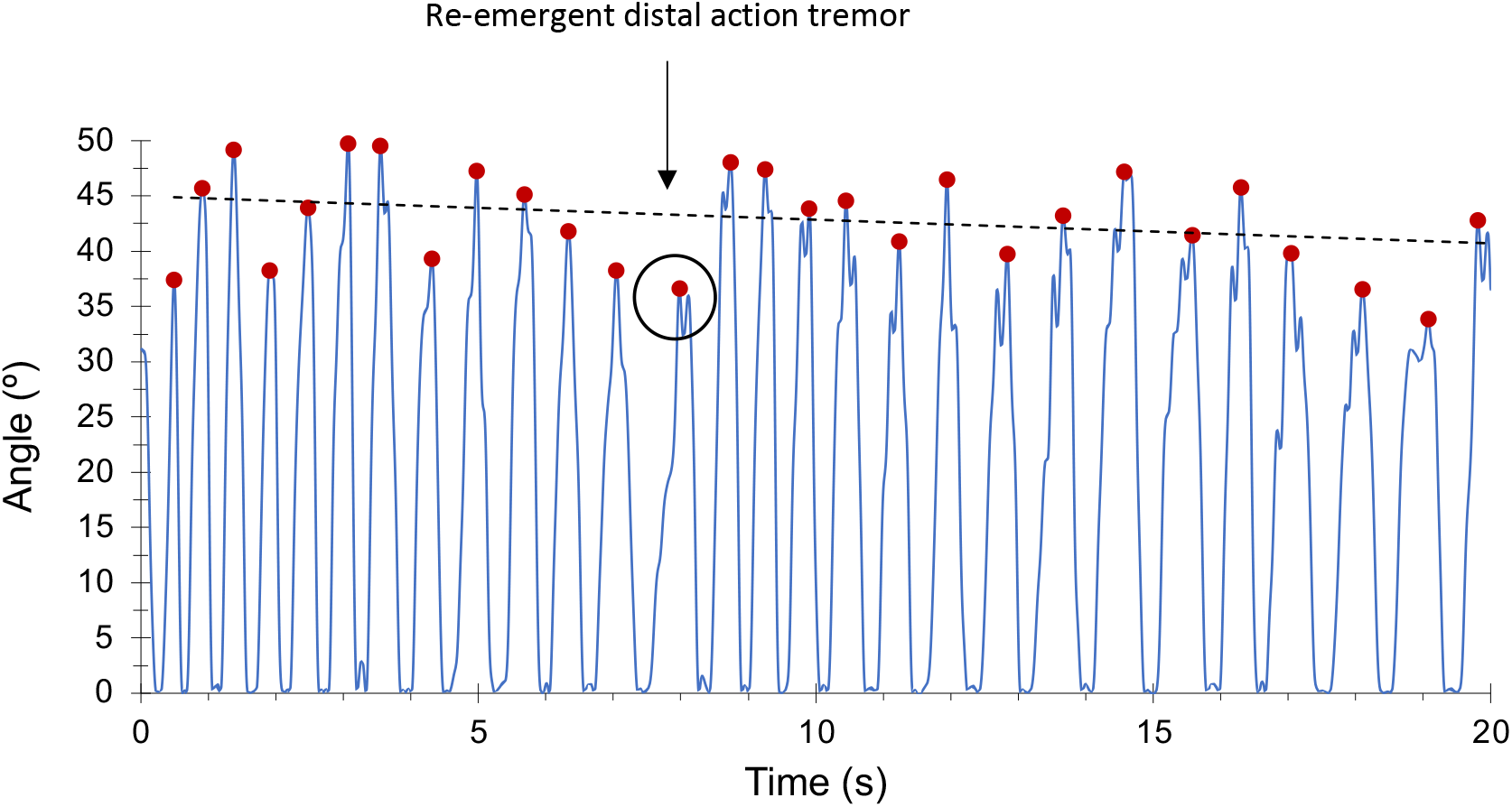
Patient with PD with index finger action tremor appearing after 10 seconds of latency (re-emergence phenomena). Only the highest peak of amplitude is selected.

**Table 5.**
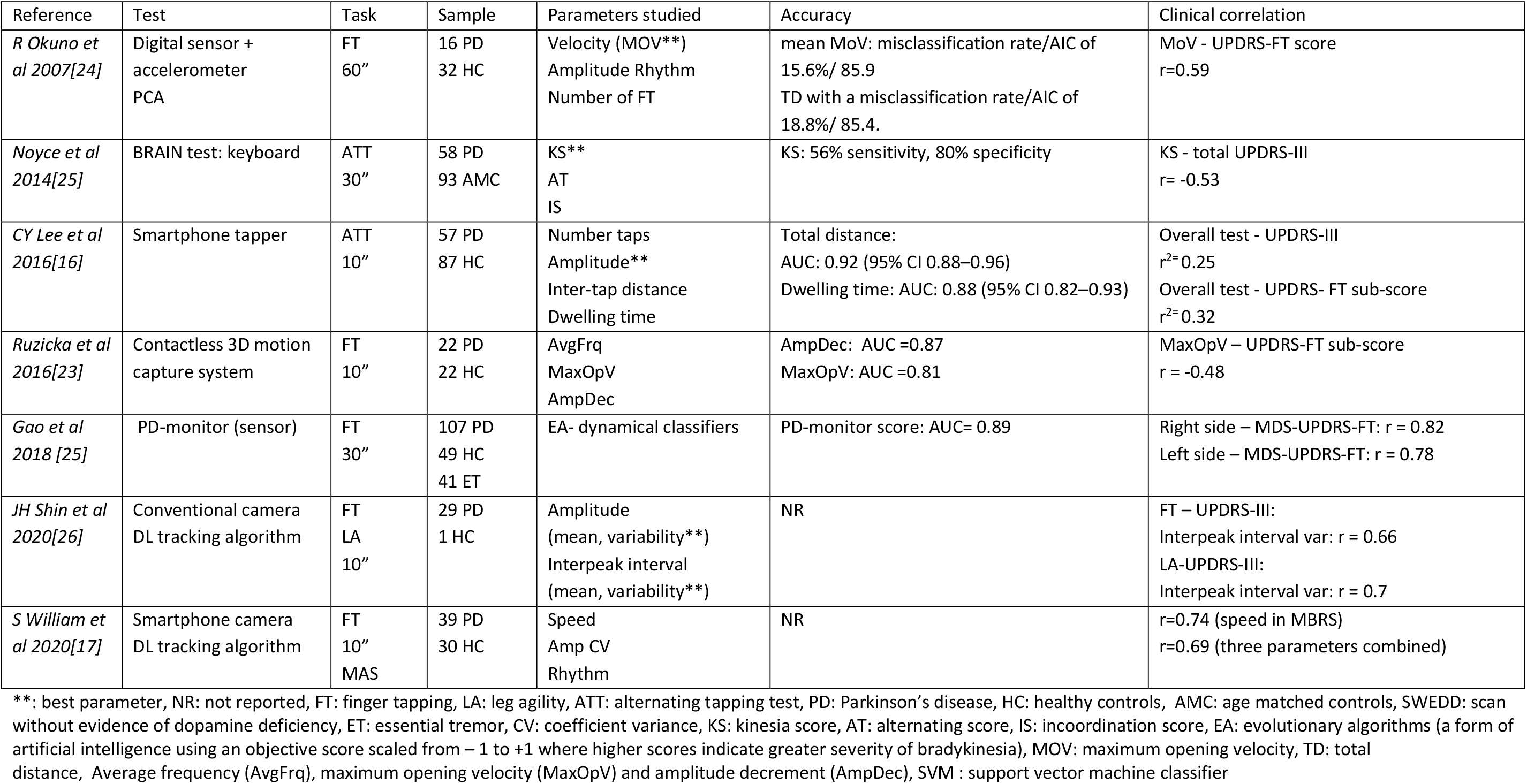
Representative literature about quantitative measures of finger movements.

| Reference | Test | Task | Sample | Parameters studied | Accuracy | Clinical correlation |
| --- | --- | --- | --- | --- | --- | --- |
| *R Okuno et al 2007[24]* | Digital sensor + accelerometer<br>PCA | FT<br>60" | 16 PD<br>32 HC | Velocity (MOV**)<br>Amplitude Rhythm<br>Number of FT | mean MoV: misclassification rate/AIC of 15.6%/ 85.9<br>TD with a misclassification rate/AIC of 18.8%/ 85.4. | MoV - UPDRS-FT score<br>$r=0.59$ |
| *Noyce et al 2014[25]* | BRAIN test: keyboard | ATT<br>30" | 58 PD<br>93 AMC | KS**<br>AT<br>IS | KS: 56% sensitivity, 80% specificity | KS - total UPDRS-III<br>$r= -0.53$ |
| *CY Lee et al 2016[16]* | Smartphone tapper | ATT<br>10" | 57 PD<br>87 HC | Number taps<br>Amplitude**<br>Inter-tap distance<br>Dwelling time | Total distance:<br>AUC: 0.92 (95% CI 0.88–0.96)<br>Dwelling time: AUC: 0.88 (95% CI 0.82–0.93) | Overall test - UPDRS-III<br>$r^2= 0.25$<br>Overall test - UPDRS- FT sub-score<br>$r^2= 0.32$ |
| *Ruzicka et al 2016[23]* | Contactless 3D motion capture system | FT<br>10" | 22 PD<br>22 HC | AvgFrq<br>MaxOpV<br>AmpDec | AmpDec: AUC =0.87<br>MaxOpV: AUC =0.81 | MaxOpV – UPDRS-FT sub-score<br>$r = -0.48$ |
| *Gao et al 2018 [25]* | PD-monitor (sensor) | FT<br>30" | 107 PD<br>49 HC<br>41 ET | EA- dynamical classifiers | PD-monitor score: AUC= 0.89 | Right side – MDS-UPDRS-FT: $r = 0.82$<br>Left side – MDS-UPDRS-FT: $r = 0.78$ |
| *JH Shin et al 2020[26]* | Conventional camera<br>DL tracking algorithm | FT<br>LA<br>10" | 29 PD<br>1 HC | Amplitude<br>(mean, variability**)<br>Interpeak interval<br>(mean, variability**) | NR | FT – UPDRS-III:<br>Interpeak interval var: $r = 0.66$<br>LA-UPDRS-III:<br>Interpeak interval var: $r = 0.7$ |
| *S William et al 2020[17]* | Smartphone camera<br>DL tracking algorithm | FT<br>10"<br>MAS | 39 PD<br>30 HC | Speed<br>Amp CV<br>Rhythm | NR | $r=0.74$ (speed in MBRS)<br>$r=0.69$ (three parameters combined) |

\*\* : best parameter, NR: not reported, FT: finger tapping, LA: leg agility, ATT: alternating tapping test, PD: Parkinson's disease, HC: healthy controls, AMC: age matched controls, SWEDD: scan without evidence of dopamine deficiency, ET: essential tremor, CV: coefficient variance, KS: kinesia score, AT: alternating score, IS: incoordination score, EA: evolutionary algorithms (a form of artificial intelligence using an objective score scaled from – 1 to +1 where higher scores indicate greater severity of bradykinesia), MOV: maximum opening velocity, TD: total distance, Average frequency (AvgFrq), maximum opening velocity (MaxOpV) and amplitude decrement (AmpDec), SVM : support vector machine classifier

Amplitude and velocity from tapping tasks correlated best with the MDS-UPDRS-III finger tapping sub-scores and might therefore be useful surrogate markers for assessing disease severity. It is however important to consider that two different means of data were compared, categorical (from normal to severe FT sub-score) and continuous data (SMART test parameters). One might expect a floor effect, as it can be interpreted from correlation graphs in the supplementary material (Figure 1), between lower categorical scores (slight and mild score) which continuous data might be better able to define. Although there was a moderate positive correlation with FT sub-scores, the lack of any stronger correlation suggests that the SMART test and the finger tapping sub-scores of the MDS-UPDRS-III are identifying different phenomena. Williams and colleagues carried out a project with a similar approach *[22]*. Smartphone video recordings of a 10-second finger tapping task were tracked with DeepLabCut (CNN). In this study patients had a longer disease duration (median of 4 years) and were on average 9 years older than ours. Although accuracy was not reported, the velocity parameter exhibited a greater correlation with FT-sub-score of MDS-UPDRS-III than ours (r: −0.74 vs r: −0.60). This may support the notion that the MDS-UPDRS-III is best adapted to patients with established disease rather than earlier stages *[23]*, suggesting that the findings from this study should be confirmed in people with longer disease duration. In line with the previous study, Schneider and colleagues studied patients with PD (around 4 years of disease duration). Patients were tested using a semiquantitative scale integrated in a motor battery which covered arm swing assessment, single finger tremor, number of finger taps, and handwriting analysis. Whilst the number of repetitive fingers taps per minute was similar between groups, ‘fatigability’ (decrement of amplitude) was more evident among patients. Although the findings were descriptive, they believe that their battery was capable of detecting early subtle motor markers that might be missed by the UPDRS-III *[24]*.

Slow motion tracking of repetitive finger tapping may help to understand how fast, fluid, and erratic normal voluntary movements are. Beyond the decrement of amplitude and frequency, defined as ‘*sequence effect’* in bradykinesia, non-linear patterns are seen among patients and controls which make it more difficult to establish cut-offs for normal. It is important to consider that clinical scales are semi-quantitative and semi-objective, and they are prone to individual bias which increases inter- and intra-rater variability *[23]*. To be of practical value, technology should exceed the performance of “Gold Standard” clinical scales or at least be more efficient.

A study conducted in 384 patients at an early stage of PD (2 or less years from diagnosis), highlighted that limitation of the MDS-UPDRS-III in early PD. The motor impact shown by MDS-UPDRS-II (capturing motor experiences) did not correlate well with motor severity of motor signs detected by MDS-UPDRS-III, especially in those with very mild degrees of severity *[25]*. A marked floor effect (large concentration of clinical phenotypes near the lower limit) of clinical appeared to be the key reason for that gap. The authors concluded that MDS-UPDRS-III had clinimetric limitations which could reduce its accuracy in early disease. In contrast, technology could potentially overcome this limitation. Gao and collaborators designed a sensor device able to assess finger tapping and explore whether it could be used to identify early stages of PD and correlate with disease progression *[26]*. Readings from the sensors were analysed by using evolutionary algorithms which are a form of artificial intelligence designed to create classifiers of patterns of movement *[27]*. Their tool reached a high accuracy (≥89.7%) for detecting different severity degrees of bradykinesia. Moreover, it could discriminate early stages of PD with AUC of 0.899. These findings should encourage further research to focus on meticulous detection methods of motor dysfunction throughout the disease course, including the prodromal phase of PD. In fact, a recent review gave evidence about the potential role of video-based artificial intelligence in PD diagnosis and monitoring which could be particularly useful when classification involves complex and dynamical patterns of movement *[28]*.

Our study is the first to use a technology-based tool to look for subtle motor features in idiopathic anosmia. Although our findings remain exploratory and warrant further investigation in a larger sample, the SMART test appeared able to detect subtle changes in anosmia group whilst the finger-tapping sub-score of the MDS-UPDRS-III was less able to identify such discrepancies (6 out 9 patients had normal finger tapping sub-scores). In our sample, finger-tapping performance in individuals with anosmia clearly differed from the control group (Figure 4). Anosmia is a prodromal marker of future PD risk *[29]*. The Health, Aging and Body Composition study showed the hazard ratio for PD over 10 years of follow up to be 4.8 for subsequent PD diagnosis *[30]*. Another large population-based cohort, the PRIPS study, reported a relative risk ratio of 6.5 in participants with reduced sense of smell after 3 years follow-up *[31]*. Most studies of idiopathic anosmia did not find detectable motor dysfunction using the MDS-UPDRS-III *[32–34]*. One longitudinal study showed that whereas subjects with hyposmia did not have worse UPDRS-III scores than individuals with a normal sense of smell, a greater proportion had abnormalities on dopamine transporter SPECT (11% vs. 1%) *[33]*. One systematic review and meta-analysis suggested that anosmia was associated with a 3.84-fold risk of developing PD *[35]* and the MDS Criteria for Prodromal PD show that, based on seven prospective studies, objective smell loss has a positive likelihood ratio of 4.0 *[7]*. Based on these findings, the presence of motor features in some patients with anosmia might be expected. The fact that UPDRS-III is often normal in patients with anosmia suggests that other assessments adapted for early stages of PD are needed *[36]*.

The SMART test offers several advantages. It is a sensor-free tool; therefore, it does not interfere with the natural range of movement. It is inexpensive with a smartphone camera only being required which can potentially make it applicable in larger scale studies. However, it also entails several methodological and data processing limitations.

In terms of methodology, one limitation might be that the exclusion of controls scoring more than 6 in the MDS-UPDRS-III (cut off for subthreshold parkinsonism) may have contributed to artificially increasing test accuracy. Another limitation to consider is that by asking to not rotate the hand which was done to capture the real angle we might have prevented patients to adopt a dystonic hand posture. It would be particular important in patients exhibiting action tremor since a possible co-existence of dystonic action tremor could be expected, especially in early diagnosed patients. Finally, although we tested for a longer period of time than it is recommended by the MDS-UPDRS-III (10 seconds), we should consider testing for longer than 20 seconds, especially in patients at earlier stages.

Moving to data processing limitations, we derived relatively simple summary statistics from the derived time series, and it may be using other techniques based on the frequency domain that capture beat-to-beat variation may be more sensitive, as demonstrated by Biase and colleagues with the tremor stability index *[37]*. However, the aim of this work was to provide proof of concept, that motion capture using a smart phone could provide metrics sensitive to changes in early PD. There are a large number of non-linear, time-series metrics, and this question will be the focus of future work. Although we used a simple, threshold-based method, for discriminating PD from controls, we acknowledge that there are other approaches based on machine learning that may be able to leverage the whole time-series, or indeed the raw video footage, and ultimately prove more accurate. However, in this work we sought to derive quantitative metrics from video footage, given these measures have much broader utility beyond mere categorical diagnostics (e.g. treatment biomarkers). Finally, we did not find a difference between ‘on’ and ‘off’ stages whereas MDS-UPDRS-III did find a 40% change. A reasonable explanation for that would be that MDS-UPDRS-III covers the ‘whole picture’ (walking, facial expression, rigidity, etc) whereas finger tapping only assesses distal bradykinesia.

## CONCLUSIONS

The SMART test provides objective evidence of motor dysfunction in PD with velocity being the best parameter to differentiate recently diagnosed PD cases from controls. Individuals with idiopathic anosmia exhibited abnormal patterns of movement supporting the idea of anosmia being part of the prodromal phase of PD.

## Data Availability

The authors confirm that the data supporting the findings of this study are available within the article [and/or] its supplementary materials.

## ETHICAL COMPLIANCE STATEMENT

We confirm that we have read the Journal’s position on issues involved in ethical publication and affirm that this work is consistent with those guidelines. The authors confirm that all participants gave verbal and written consent for this work. Ethics approval was granted by the PREDICT-PD study was approved by Central London Research Committee 3 (reference number 10/H0716/85). The qMAP-PD study has full NHS ethical approval (Fulham Research Ethics Committee, 18/LO/1229).

## TABLES AND FIGURES

**Table 1** Demographic and clinical data.

**Table 2** Test parameter comparison (PD vs Controls)

**Table 3.** ROC analysis (PD vs Controls).

**Table 4.** Test parameter comparison (Anosmia vs Controls).

**Table 5.** Representative literature about quantitative measures of finger movements.

**Figure 1.** Hand detection: 8 key landmarks across the first and the second finger (red). Angle between 1,4,8 key landmarks (black). Extrapolated amplitude between point 1 and 8 (blue).

**Figure 2.** Receiver operator characteristic (ROC) curves for the best parameter combination to distinguish patients with PD and controls. A) Velocity and CV frequency (AUC 0.83; 95% CI 0.72 to 0.95), B) Velocity and frequency slope (AUC 0.88, 95% CI 0.78 to 0 0.97), C) CV velocity and frequency slope (AUC 0.85; 95% CI 0.74 to 0.95).

**Figure 3.** Representative examples of one control (A), PD (B), and anosmia (C). Finger tapping sub-scores (MDS-UPDRS-III): 0 (control and anosmia), 1 (PD). Control: constant amplitude and frequency. PD amplitude with ‘*burst phenomena*’and frequency decrease. Anosmia: smaller amplitude and frequency. compared with control.

**Figure 4.** Patient with PD with index finger action tremor appearing after 10 seconds of latency (re-emergence phenomena). Only the highest peak of amplitude is selected.

