## Supplemental data for "Slow Motion Analysis of Repetitive Tapping (SMART) test: measuring bradykinesia in recently diagnosed Parkinson’s disease and idiopathic anosmia"

**Supplementary material**

| **Table 1. The Movement Disorder Society revision of the Unified Parkinson's Disease Rating Scale (MDS-UPDRS) Item 3.4 (Finger Tapping). MDS-UPDRS grade** | |
| --- | --- |
| **Score** | **Description** |
| 0 -Normal | - No problems |
| 1 -Slight | - Any of the following: a) the regular rhythm is broken with one or two interruptions or hesitations of the tapping movement; b) slight slowing; c) the amplitude decrements near the end of the 10 taps. |
| 2 -Mild | - Any of the following: a) 3 to 5 interruptions during tapping; b) mild slowing; c) the amplitude decrements midway in the 10-tap sequence. |
| 3 -Moderate | - Any of the following: a) more than 5 interruptions during tapping or at least one longer arrest (freeze) in ongoing movement; b) moderate slowing; c) the amplitude decrements starting after the 1st tap |
| 4 -Severe | - Cannot or can only barely perform the task because of slowing, interruptions, or decrements. |

Each hand is tested separately. The patient is instructed to tap the index finger on the thumb “as quickly and as big as possible”

| **Table 2. Correlation between SMART test parameters and MDS-UPDRS-III FT sub-scores** | | |
| --- | --- | --- |
| PD cases (n=24)* | **Spearman correlation with FT sub-score** | ***p* value** |
| **Amplitude** Mean | -0.49 (-0.81 to -0.17) | 0.003 |
| CV | -0.07 (-0.36 to 0.49) | 0.758 |
| Slope | 0.44 (0.07 to 0.81) | 0.018 |
| **Frequency** Mean | 0.175 (-0.24 to 0.59) | 0.414 |
| CV | 0.25 (-0.16 to 0.66) | 0.239 |
| Slope | -0.16 (-0.58 to 0.25) | 0.443 |
| **Velocity** Mean | -0.43 (-0.78 to -0.08) | 0.016 |
| CV | 0.44 (0.09 to 0.79) | 0.013 |
| Slope | 0.34 (0.05 to 0.74) | 0.087 |
| All parameters presented with 95% coefficient interval (CI). FT: Finger tapping UPDRS: Unified Parkinson’s disease rating scale, CV: coefficent variation. * 2 cases scored 0 and 3. They were excluded since they might influence the regression. | | |

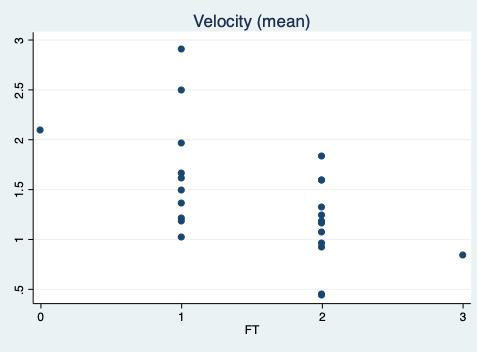

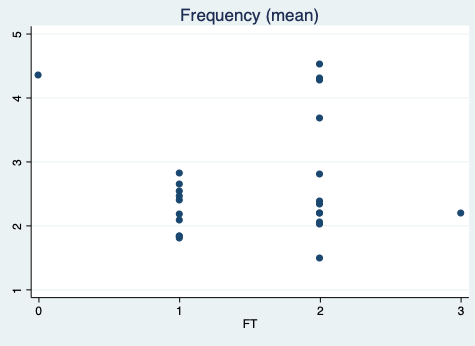

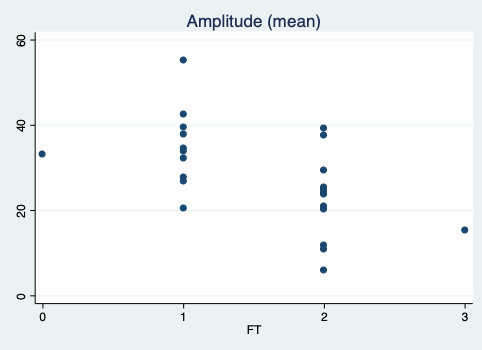

Figure 1. Correlation between SMART test parameters and finger-tapping sub-score from MDS-UPDRS-III: 0 (normal), 1 (slight), 2 (mild), 3 (moderate). There were no PD cases scoring 4 (severe). Evidence of floor effect (wide range of SMART test performance) between score 1 and 2. NOTE: detailed scoring definition is in supplementary material.

C)

r= -0.43

*p*= 0.016

r= 0.18

*p*= 0.414

r= -0.49

*p*= 0.003

B)

A)
